# Better Strategies for Containing COVID-19 Epidemics–A Study of 25 Countries via an Extended SEIR Model

**DOI:** 10.1101/2020.04.27.20081232

**Authors:** Jia Gu, Han Yan, Yaxuan Huang, Yuru Zhu, Haoxuan Sun, Xinyu Zhang, Yuqing Wang, Yumou Qiu, Song Xi Chen

## Abstract

We evaluate the effectiveness of COVID-19 control strategies of 25 countries which have endured more than four weeks of community infections. With an extended SEIR model that allows infections in both the exposed and infected states, the key epidemic parameters are estimated from each country’s data, which facilitate the evaluation and cross-country comparison. It is found quicker control measures significantly reduce the average reproduction numbers and shorten the time length to infection peaks. If the swift control measures of Korea and China were implemented, average reductions of 88% in the confirmed cases and 80% in deaths would had been attained for the other 23 countries from start to April 10. Effects of earlier or delayed interventions in the US and the UK are experimented which show at least 75% (29%) less infections and deaths can be attained for the US (the UK) under a Five-Day Earlier experiment. The impacts of two removal regimes (Korea and Italy) on the total infection and death tolls on the other countries are compared with the natural forecast ones, which suggest there are still ample opportunity for countries to reduce the final death numbers by improving the removal process.

## Introduction

The Corona Virus Disease 2019 (COVID-19) has spread to the world causing a pandemic with more than 2 millions infections and more than 148 thousands deaths world wide (*1*) on April 19th, 2020. There have been studies for the effects of early control measures taken in China: (*2–4*) on public health interventions and control strategies on Wuhan’s outbreaks; (*5*) on Wuhan travel ban on the spread of COVID-19 inside China and (*6*) on both the domestic and international implications of Wuhan travel ban; (*7*) on the transmissibility and severity in mainland Chinese locations outside Hubei. Three months after Wuhan being sealed off, there are more than 25 countries which have endured at least four weeks of community infections with good amount of epidemic data accrued (*8–10*). Given the unprecedented pressure on nations’ healthcare systems and the deaths so far, there is an urgent need to learn from these countries’ paths when other countries plan to counter the epidemic.

Our evaluation is based an extended SEIR model (*11–15*) with time varying coefficient (vSEIdR model) that permits infections in both the Infected state and the Exposed state to reflect the COVID-19 clinical reality that more infections are made before cases being diagnosed (*16*). We provide frequentist estimates to the infection, diagnosis and removal rates, and the effective reproduction number *R_t_* which may be time-varying. The latter allows capturing the changing dynamics in the spread of the virus and to assess the effects of different policy scenarios.

Twenty-one out of the twenty five countries have reached the turning point of infection defined as the first day that *R_t_* < 1 since the start of community infection. The average time to the turning point is 34.1 days (SE 2.27), but among the 10 countries which took action to reduce person to person contacts sooner (see Table A1 for specifics), the average time to the turning points is 28.7 days (3.5), which is significantly (p-value 0.026) lower than the later action group (mean 37.8, SE 2.7). Taking early control measures is also effective in reducing the infection rates leading to suppressing the effective reproduction numbers *R_t_* by 0.819 (p-value 0.007) in weeks 2–4 after the start of the epidemic between the quick and slow action groups of countries.

China (excluding Hubei Province) and Korea are found to be the most effective in bring down the reproduction of COVID-19 in the first four weeks of community infections (Table 1 and Figure 1). The benefits of acting early in reducing both the infection size and total deaths are demonstrated by counter-factual calculations under the Korea and China scenarios. Figure 2 shows reductions of 1.20 million (1.26 millions) infected cases and 75,105 (79,543) deaths among the 23 countries would be made if Korea (China)’s daily reduction percentage in the infection rates are adopted from Day 8 of each country’s start of community infection to April 10, while maintaining each country’s removal and diagnostic rates. These numbers mount to 86% (90%) and 78% (82%) of the total infected cases and deaths of the 23 countries on April 10th, respectively.

**Table 1:** Weekly averages of the estimated reproduction numbers *R_t_* (W1-W4) of 25 countries over the four weeks from their respective start date of community transmission (DCT). Countries are ranked based on the average *R_t_* over the four weeks (4W-Ave). China refers to the provinces excluding Hubei province. The 95% confidence intervals for *R*_0_ are available in Table S1 in SM.

| | Country | DCT | $R_0$ | W1 | W2 | W3 | W4 | 4W-Ave |
| --- | --- | --- | --- | --- | --- | --- | --- | --- |
| 1 | China | 01-23 | 4.78 | 2.79 | 1.09 | 0.24 | 0.00 | 1.03(0.87-1.20) |
| 2 | Korea | 02-17 | 5.56 | 3.68 | 1.72 | 0.58 | 0.20 | 1.54(1.43-1.66) |
| 3 | Denmark | 03-03 | 6.26 | 3.42 | 1.10 | 1.40 | 1.68 | 1.90(1.09-2.39) |
| 4 | Austria | 03-07 | 3.68 | 3.15 | 2.40 | 1.45 | 0.60 | 1.90(1.21-2.12) |
| 5 | Singapore | 03-04 | 2.48 | 2.42 | 2.36 | 1.64 | 1.46 | 1.97(1.45-2.36) |
| 6 | Japan | 02-12 | 4.83 | 3.17 | 1.79 | 2.07 | 1.63 | 2.17(1.51-2.77) |
| 7 | Norway | 03-03 | 5.21 | 4.14 | 1.95 | 1.55 | 1.07 | 2.18(2.00-2.20) |
| 8 | Thailand | 03-07 | 4.57 | 4.29 | 2.65 | 1.37 | 0.75 | 2.27(1.84-2.49) |
| 9 | Malaysia | 02-29 | 4.46 | 3.31 | 3.09 | 1.74 | 1.20 | 2.34(1.21-2.96) |
| 10 | Switzerland | 03-01 | 3.64 | 3.46 | 3.08 | 1.89 | 1.20 | 2.41(2.21-2.53) |
| 11 | Canada | 03-07 | 3.62 | 3.31 | 2.87 | 2.17 | 1.59 | 2.48(2.20-2.61) |
| 12 | Sweden | 03-01 | 6.02 | 4.67 | 1.92 | 1.79 | 2.10 | 2.62(2.49-2.72) |
| 13 | Iran | 02-22 | 8.62 | 5.37 | 2.21 | 1.58 | 1.56 | 2.68(2.37-3.11) |
| 14 | Turkey | 03-18 | 5.54 | 4.59 | 2.60 | 1.98 | 1.66 | 2.71(2.78-2.98) |
| 15 | Portugal | 03-07 | 5.93 | 5.09 | 3.21 | 2.10 | 1.20 | 2.90(1.77-4.01) |
| 16 | Italy | 02-23 | 6.25 | 4.42 | 3.38 | 2.61 | 1.80 | 3.05(2.95-3.18) |
| 17 | Belgium | 03-04 | 4.95 | 4.42 | 3.42 | 2.79 | 1.57 | 3.05(2.79-3.30) |
| 18 | Brazil | 03-10 | 5.65 | 4.91 | 2.79 | 2.56 | 1.96 | 3.06(2.90-3.21) |
| 19 | Australia | 02-26 | 4.83 | 3.88 | 3.80 | 3.46 | 2.07 | 3.30(2.84-3.77) |
| 20 | US | 02-29 | 4.10 | 3.82 | 4.38 | 3.33 | 2.24 | 3.44(3.33-3.51) |
| 21 | Holland | 02-29 | 6.74 | 6.06 | 3.51 | 2.92 | 1.85 | 3.58(3.03-4.11) |
| 22 | Spain | 02-29 | 6.75 | 6.34 | 4.22 | 2.98 | 1.74 | 3.82(3.56-4.00) |
| 23 | France | 02-25 | 7.03 | 6.13 | 3.92 | 2.95 | 2.49 | 3.87(3.43-4.25) |
| 24 | Germany | 02-25 | 6.43 | 6.01 | 4.87 | 3.57 | 2.07 | 4.13(3.65-4.71) |
| 25 | UK | 02-25 | 7.03 | 5.86 | 4.49 | 3.77 | 3.20 | 4.33(3.82-4.43) |
| Ave |  |  | 5.40 | 4.35 | 2.91 | 2.18 | 1.56 | 2.75 |
| SE |  |  | 0.37 | 0.23 | 0.21 | 0.18 | 0.14 | 0.16 |

**Figure 1:**
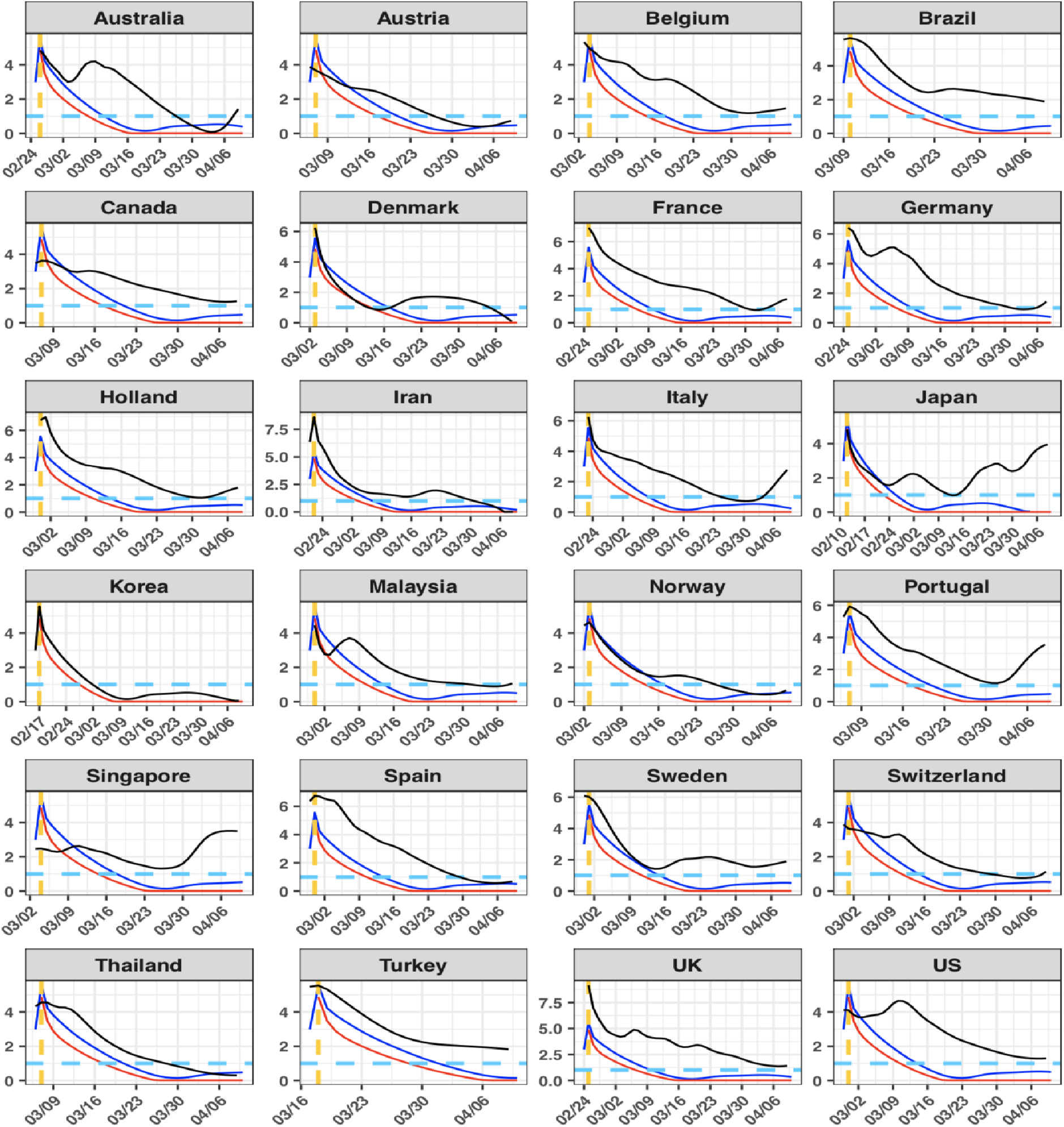
The estimated effective reproduction number *R_t_* curves (black) of the countries since their start dates of community transmission (yellow dashed vertical lines) versus those of China (red) and Korea (blue). The blue dashed line represents the critical threshold level 1.

**Figure 2:**
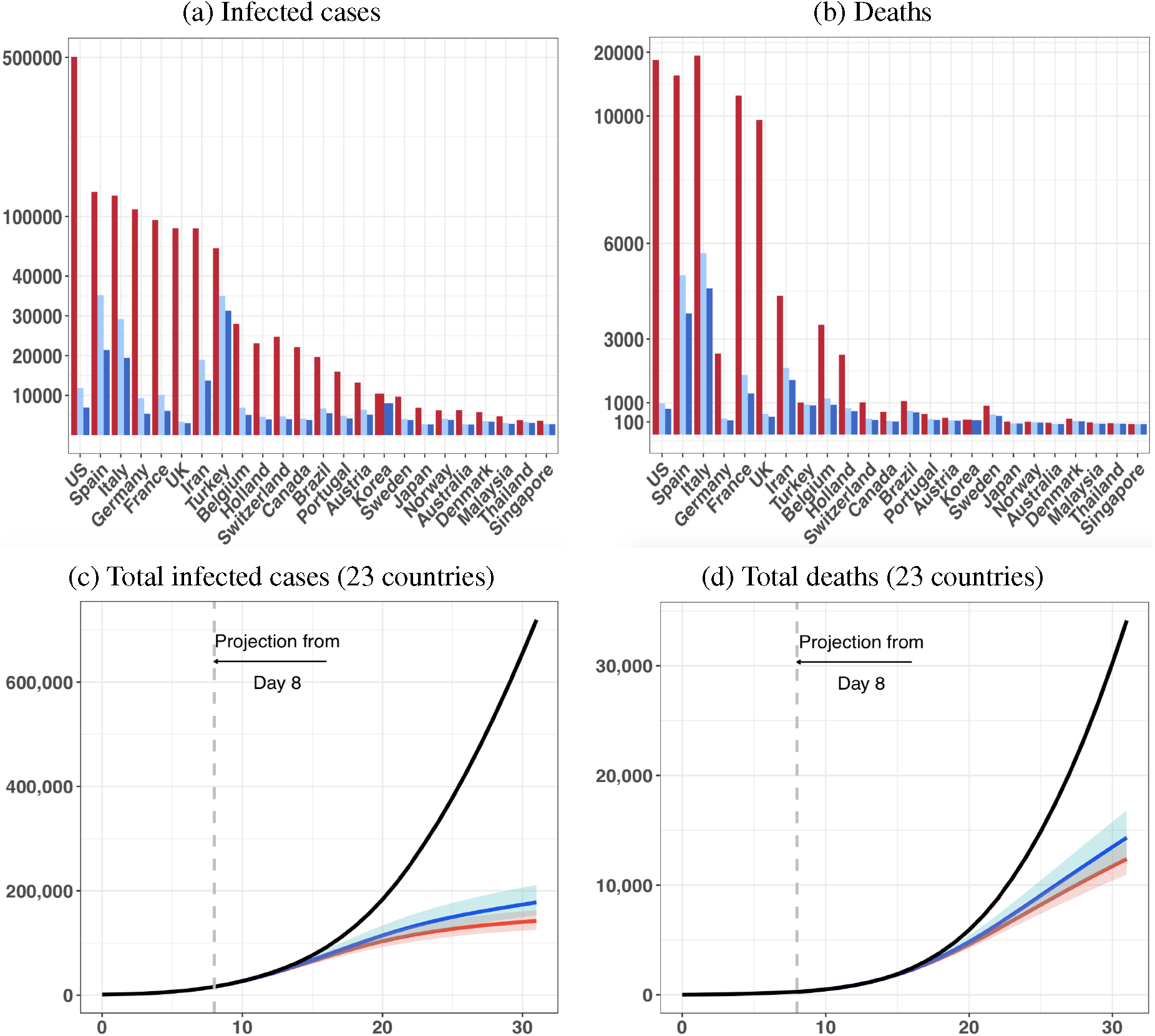
The observed numbers (red bar) of infected cases (a) and deaths (b) of the countries and the would-be ones under China (blue bar) and Korea (light blue bar)’s scenarios implemented from Day 8 of community transmission to April 10; and the observed (black) total numbers of infected cases (c) and deaths (d) of the 23 countries excluding Korea and China and the would-be totals from Day 8 to Day 31 under China (red) and Korea (blue)’s scenarios.

**Figure 3:**
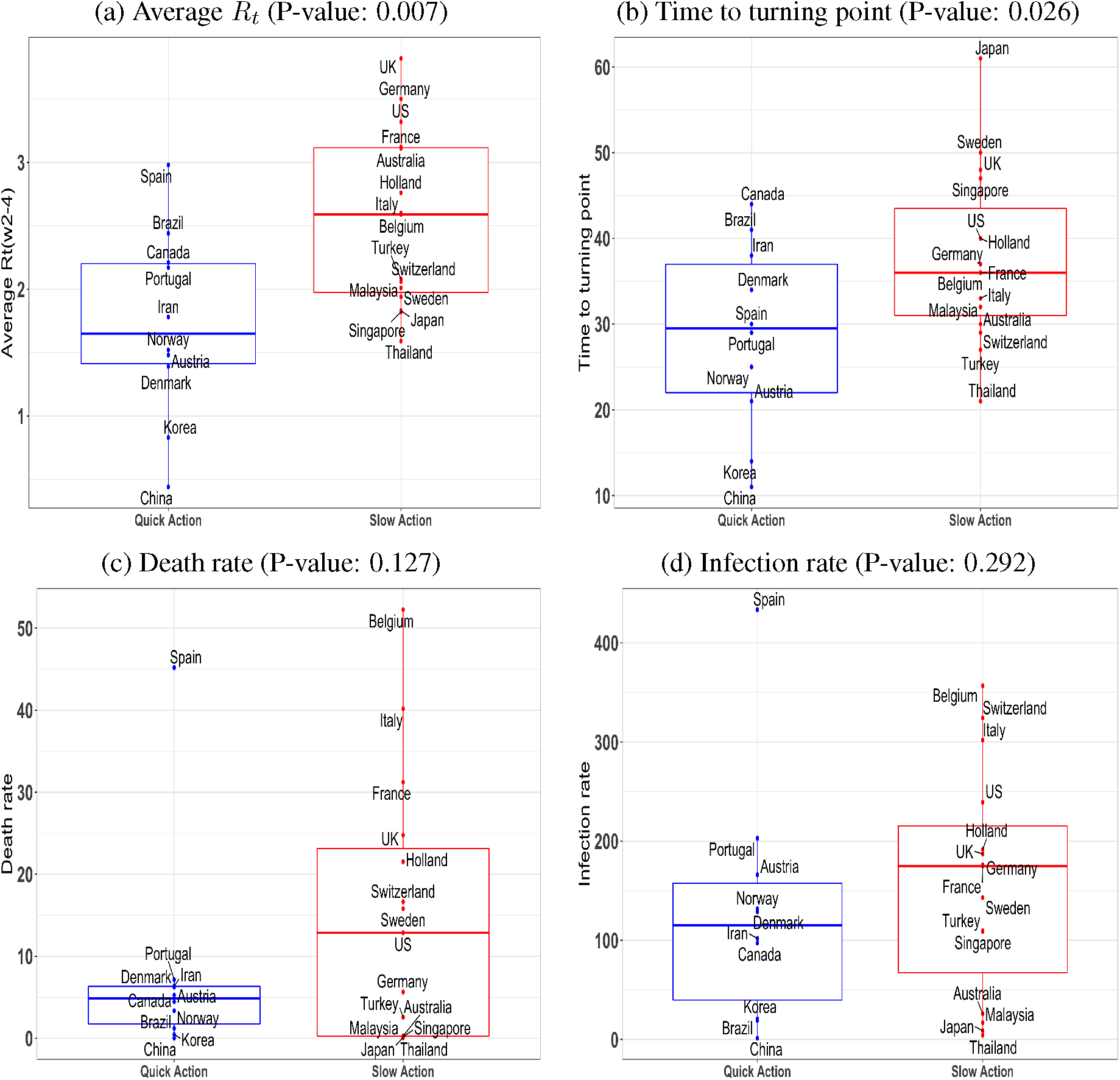
Comparative box plots for four variables between the 10 Quick Action countries (time from the start of community transmission to decisive restriction measures less than 13 days) and the 15 Slow Action countries: the average estimated reproduction number *R_t_* in weeks 2–4 since the start date of community transmission (a), the time from the start date to the turning point (first date with *R_t_ <* 1) (b), the death number per 100,000 population (c) and the infection number per 100,000 population (d). The p-values of the one-sided two-sample t-test are reported in the parentheses appeared in the subtitles.

That taking control measures sooner can significantly impact the sizes of epidemics and deaths can be shown without having to mimic Korea and China’s experiences. For the US and the UK, if policy interventions were made to ensure the declines in the *R_t_* from March 13 for the US (*17*) and March 20 for the UK (*18*) happened five days earlier (delayed), the US would have the cases and the deaths reduced (increased) by about 80% and 75% (71% and 53%), and the UK by 40% and 29% (61% and 40%), respectively, relative to the observed statistics on April 10 (Figure 4). The US and the UK experiments also inform the role played by the diagnosis rate *α* that regulates the speed of movement from the exposed state to the infected (diagnosed) state. In particular, if the UK’s low diagnosis rate (0.1) were applied to the US (0.17), both the infection cases and the deaths would be increased by 314% and 213% under the Five-Day delayed setting.

**Figure 4:**
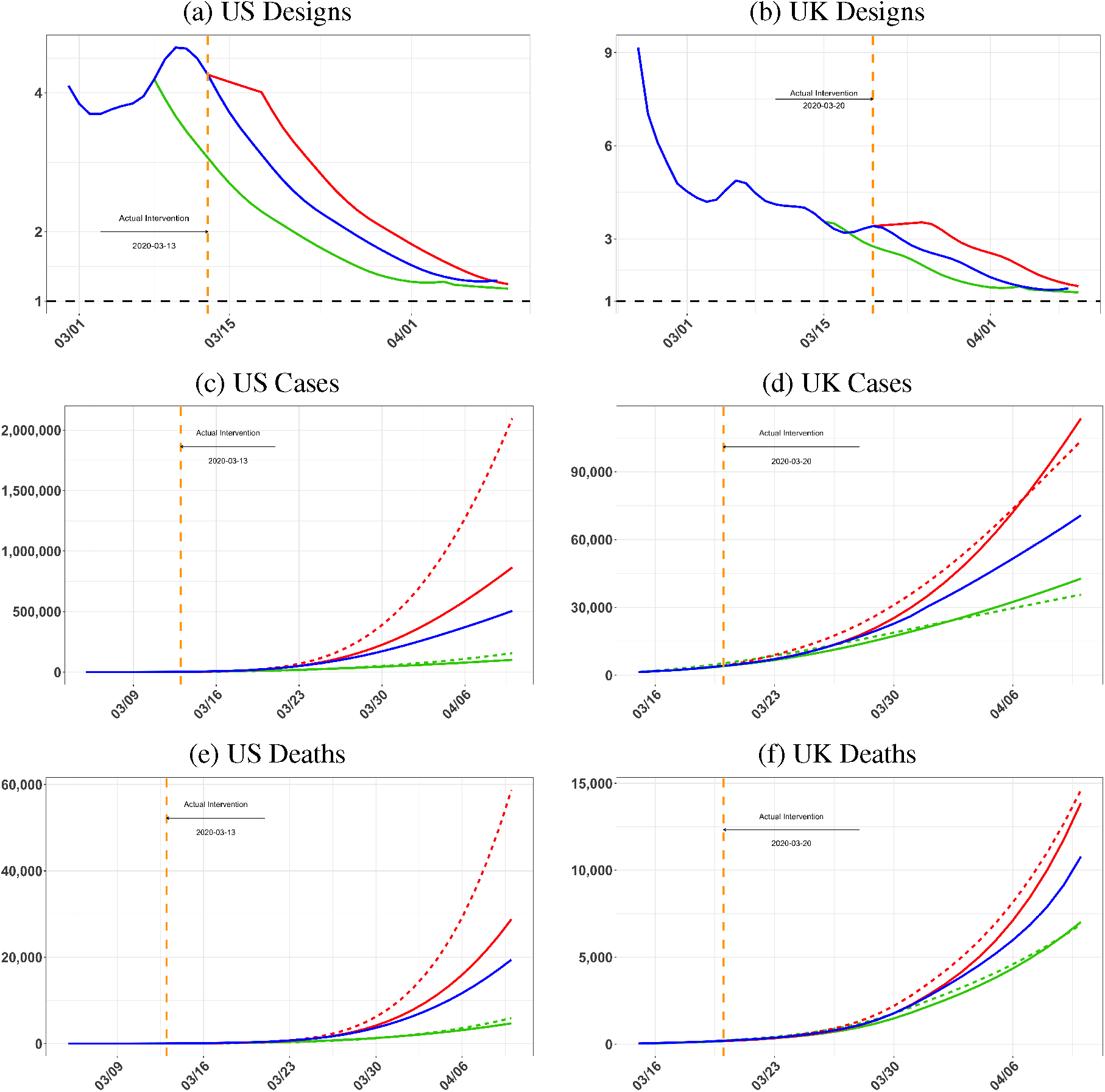
The effective reproduction number *R_t_* curves (blue lines) and the *R_t_* curves under the Five-Day Earlier (green lines) and the Five-Day Delayed (red lines) for the US (a) and the UK (b), and the infected cases (c, d) and deaths (e, f) of the observed (blue) and those generated under the Five-Day Earlier (green) and Five-Day Delayed (red) experiments. Those dashed lines are cases and deaths with the US and the UK exchanging their diagnosis rate *α*: *α*_US_ = 0.17 and *α*_UK_ = 0.1. The orange dashed lines mark the dates of the control measures of the two countries.

The study shows that the removal processes (recovery and death rates) are influential in determining the death tolls for most of the 25 countries while also so on the infection cases for countries which have not reach the turning point. Relative to each country’s so-called natural forecasting designs, the deaths would increase by 169.1% in average among the 24 countries under the Italian removal regime. However, Under the Korea’s removal rates, the deaths would decline by an average of 26.7%. Therefore, there are still ample opportunities for countries to substantially reduce the final death tolls if the removal processes can be improved.

### Effective reproduction numbers

We did not include data from Hubei province where Wuhan is the capital city for China due to incomplete observation before January 16, 2020. This actually makes the epidemic of the 25 countries more comparable as they are all started with imported cases.

We first estimate the time varying daily infection rate *β_t_* and the effective reproduction number *R_t_* under the time-varying coefficient SEIdR model for the 25 countries. The starting date of local transmission (Table 1) is identified from the estimated infection rates and by checking on each country’s early epidemic data and the date given by the WHO (*9*). Table 1 also reports the estimated reproduction numbers *R_t_* on the start date, and their weekly averages over the next four weeks. Figure 1 displays the *R_t_* curves.

The average reproductive numbers on the start, which may be viewed as the basic reproduction number *R_0_*, was 5.70 (SE: 0.37) among the 25 countries with the lower and upper 25% quartiles being 4.57 and 6.26. The 14 European countries had the highest *R*_0_, averaged at 5.82 (SE: 0.50). See Table S1 in Supplementary Material (SM) for the 95% confidence intervals of *R*_0_. As *R*_0_ is known to be subject to over-estimation (*19*) with high volatility, one may look at the average *R_t_* in the 1st week after the start of local transmission, which were averaged at 4.35 (SE: 0.23). Our estimates of *R*_0_ were higher than most of the *R*_0_s from the existing studies on COVID-19, mostly under the SEIR models, for instance 2–3 from (*20,21*) on Wuhan and 3.15 (3.04–3.26) in (*5*). Our estimate of *R*_0_ for non-Hubei China (4.78) was closer to (*22*) (*R*_0_ = 5.7 for Wuhan). A reason for our higher *R*_0_ is that the vSEIdR model allows infection prior to clinical diagnosis as reflected by the *R_t_* having two terms from equation (S.4) in SM, reflecting the infections made in the Exposed (pre-diagnosis) and Infected (Diagnosed) compartments respectively.

### Effectiveness of Korea and China’s approaches

Figure 1 shows that China and Korea’s *R_t_* curves fell sharply below the other 23 countries’ over most of the four weeks after the start dates. Their rapid decline in the reproducing power was well reflected in Table 1 as their average *R_t_* over the four weeks were 1.03 and 1.54, respectively. In contrast, 20 countries had the four week average larger than 2.0, and 10 of them more than 3.0. Not only that the absolute infectiousness of Korea and China were the lowest over the four weeks, their cumulative percentages of declines were the most as shown in Table S2. The drastic decline in the reproduction of China echos recent studies on the effectiveness of Chine’s control measures (*23, 24*). Behind Korea and China’s rapid declines in their *R_t_* were two similar but not the same strategies. China’s measures are largely based on limitation of population movement and contacts by sealing off cities and enforcing high levels of self-isolation at homes (*5,25*), which led to rapid reduction in the contact rates (*2,26*). In addition to limiting population contacts, and a quick blockade of Daegu, Korean conducted more active testing for potential infections in the population with more than half million tests being carried out in the first four weeks (*27,28*), which is an extra from China’s largely population isolation approach. In additional to the significant effects on the average reproduction numbers *R_t_* and the time to turning point by taking action to restrict population contacts earlier (Panels (a)-(b) in Figure 3), there are some evidence for lowering the death number and the infection number per 100,000 population (Panels (c)-(d) in Figure 3) by taking action earlier, although not significant due to other likely confounding factors including the varying degrees of enforcement on the control measures and differential medical capability.

### Epidemic projections under Korea and China’s scenarios

Given the effectiveness of Korea and China’s approaches in containing COVID-19, we generate scenarios for other countries that mimic Korea and China’s daily reduction percentages in the infection rates from the 8th day since the start of local transmission, while keeping their diagnosis rate, recovery and death rates intact. The generated hypothetical infection rates after Day 8 (shown in Figures S1 and S2 in SM) create scenarios for other countries should the control measures of Korea and China were implemented.

Comparing to the actual cases observed up to April 10, Figure 2 shows that 1.20 (1.26) millions total reductions in the confirmed cases and 75,105 (79,543) reduction in deaths for the 23 countries under the Korea’s (China’s) scenario, mounting to 88% and 80% reductions of the confirmed cases and deaths in average, respectively (more details in Table S3–S5). The reductions in the cases and deaths would be phenomenal for US, UK, Germany, Australia, Japan and France, attaining more than 89% reduction in the confirmed cases and 74% reduction in deaths under Korea’s scenario, if control measures were implemented earlier which would lead to the reductions in the infectious rates from Day 8 of community transmission. And those under China’s scenario would be about a few percentage points more.

### Evaluation on the US and the UK

We consider two policy intervention experiments specific to the US and the UK, which represent early and delayed implementation of counter-COVID-19 measures. March 13 and March 20 represented the dates of firm policy measures by the US and the UK governments, respectively, when the US declared the national emergency and the UK started to close schools and public facilities. It appears from Figure 4 that sustained declining trend of *R_t_* was established after March 13 and March 20 for the US and the UK, respectively. The experimental design of Five-Day Earlier intervention would mean the *R_t_* curves start to decline from March 8 for the US and March 15 for the UK at the actual daily declines rates from March 13 and 20, respectively. The Five-Day Delayed experiments is to mimic later actions that would delay the decline in *R_t_* from March 13 and 20 for five days to March 18 and 25, respectively. Panels (a)-(b) of Figure 4 display the actual *R_t_* curves with the hypothetical ones under the two designs.

The total numbers of confirmed cases and death cases under the two experiments are reported in Figure 4 (c)–(f) (Table S10 for specific numbers) along with the observed statistics up to April 10. Our result shows that by acting earlier both countries would see substantial reductions in the total number of infected cases and deaths: the US would see the cases and the deaths reduced by about 80% and 75% respectively; and the UK by 40% and 29% respectively. In contrast, under the Five-Day Delayed postulations, the US would see the cases and deaths increase by 71% and 53%, and the UK by 61% and 40%, respectively.

The above results indicate that the US is more responsive to the intervention than the UK, as the earlier (delayed) intervention would reduce (increase) more infection cases and deaths than the UK. This can be explained in several aspects. First, the absolute decline of US’s *R_t_* is 2.23, more than the UK’s 1.75 over the two weeks from March 13 and March 20, although the relative declines were comparable at 52% and 51%, respectively. Second, the US implemented the intervention relatively earlier as March 13 was the 14th day after the US’s start date while March 20 was the 24th days for the UK. Thus, the US would have more time to amplify the effect of the intervention.

A less obvious reason for the differential sensitivity lays in the estimated diagnosed rates: 0.17 for the US and 0.1 for the UK, implying the UK having longer time for diagnosing the infected than the US. The larger diagnosis rate for the US means quicker turn-over time from expose to diagnosis, which would bring early peak time for the confirmed infected cases *I*(*t*) and reduce the size of infections and hence the death number. Our result shows the numbers of cases and deaths for the US with the UK’s diagnosis rate (0.10) would make the US cases amplified by more than 243 % and the deaths by 171% under Five-Day Delay design. For the UK with the US’s higher diagnosis rate (0.17), there would be a further 10% reduction in the number of confirmed cases under 5-Day Early design, where the impacts on the death was not significant. A high diagnosis rate is part of the Korea’s counter COVID-19 strategy.

### Forecasting and projections

We consider future courses of the COVID-19 epidemic by forecasting the sizes of total infections, deaths and the peak active infection number of a country. The forecasting requires first projecting the key parameters of the epidemic processes, which we do under two designs (Natural Designs 1 and 2) for each country. The two designs impose two minimum recovery rates: *γ_r_*_,min_ = 1/17.5 based on the clinical results (*16,29, 30*) and *γ_r_*,_min_ = 1/28 with a two week transitional period, as some countries (the UK, Holland, Brazil, Norway) have severe underreporting for the recovered (see the unseemly high proportion of death in the total removal rate in Table S6 in SM), while leaving the infection, diagnosis and removal rate due to death naturally determined by each country’s epidemic forcing. We also conduct parallel projections by replacing the recovery rate (*γ_r,t_*) and the removal rates due to death (*γ_d,t_*) by those of Korea and Italy, respectively, to generate comparative results. Korea’s latest death rate *γ_d,t_* = 0.001 which is about one fifth of Italy’s (0.0050) while its total removal rate (0.0412) is more than twice of Italy’s (0.0185). Thus, the Korea removal situation represents a more effective treatment, faster recovery with less death regime, while Italy’s is of a much overwhelmed system with relatively high *γ_d,t_* and low *γ_r,t_*.

The projected total deaths vary substantially to the four designs, especially to the Italy and Korea removal regimes, relative to the two Natural designs as shown in Figure 5. With the Italian removal rates, the deaths would increase by 169.1% in average among the 24 countries, and 11 countries would increase by more than 200%. Under the Korea’s removal rates, the deaths would reduce by 26.7% in average, and for five countries, the US, the UK, Belgium, Sweden and Canada, the deaths would be more than halved, as compared to the predicted casualties under the Natural Designs. In contrast to the deaths, the predicted total infected cases (Figure 6) are less responsive to the designs, especially for countries which have had their *R_t_* < 1 on April 19th as the low *R_t_*s cushion much of the design differences for the projected final cases. It is also due to the four designs all used the same infection and diagnosis rates. The differences in the numbers of infected cases and deaths between the two Natural Designs are not much for almost all countries, although Design 1 (*γ_r_*,_min_ = 1/17.5) leads to less infections and deaths in general for countries whose recovery rate has not reached 1/17.5 benefited by a shorter infectious period.

**Figure 5:**
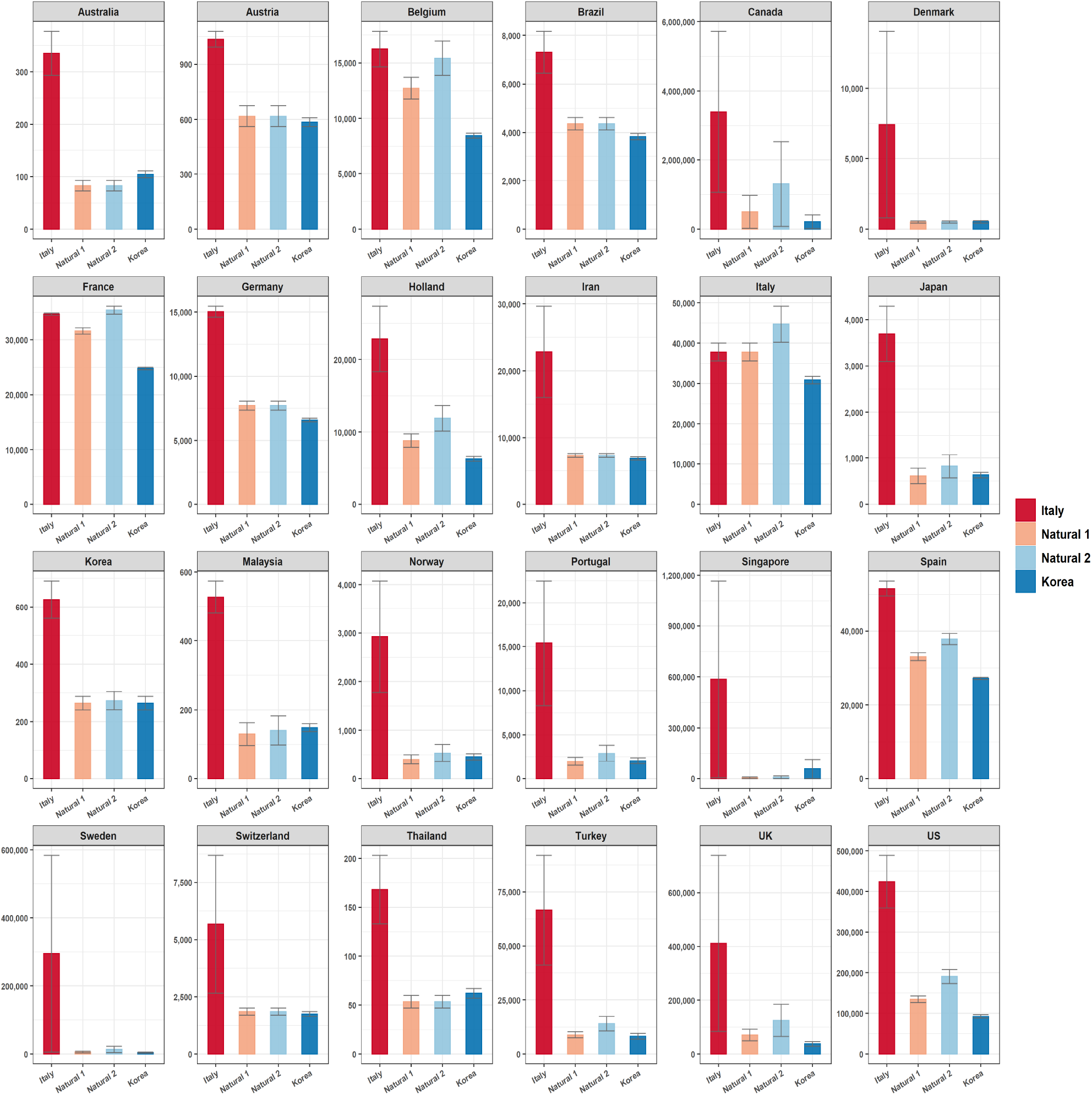
Projected total deaths and the 95% prediction intervals under the two Natural Designs of respective countries, and Korea (blue) and Italy (red)’s recovery and death rates. Natural Designs 1 and 2 impose a minimum level of 1/17.5 (orange) and 1/28 (light blue) on the recovery rate of each country, respectively. The corresponding numerical results are provided in Table S8 of SM. The data used for the forecasting are up to April 20.

**Figure 6:**
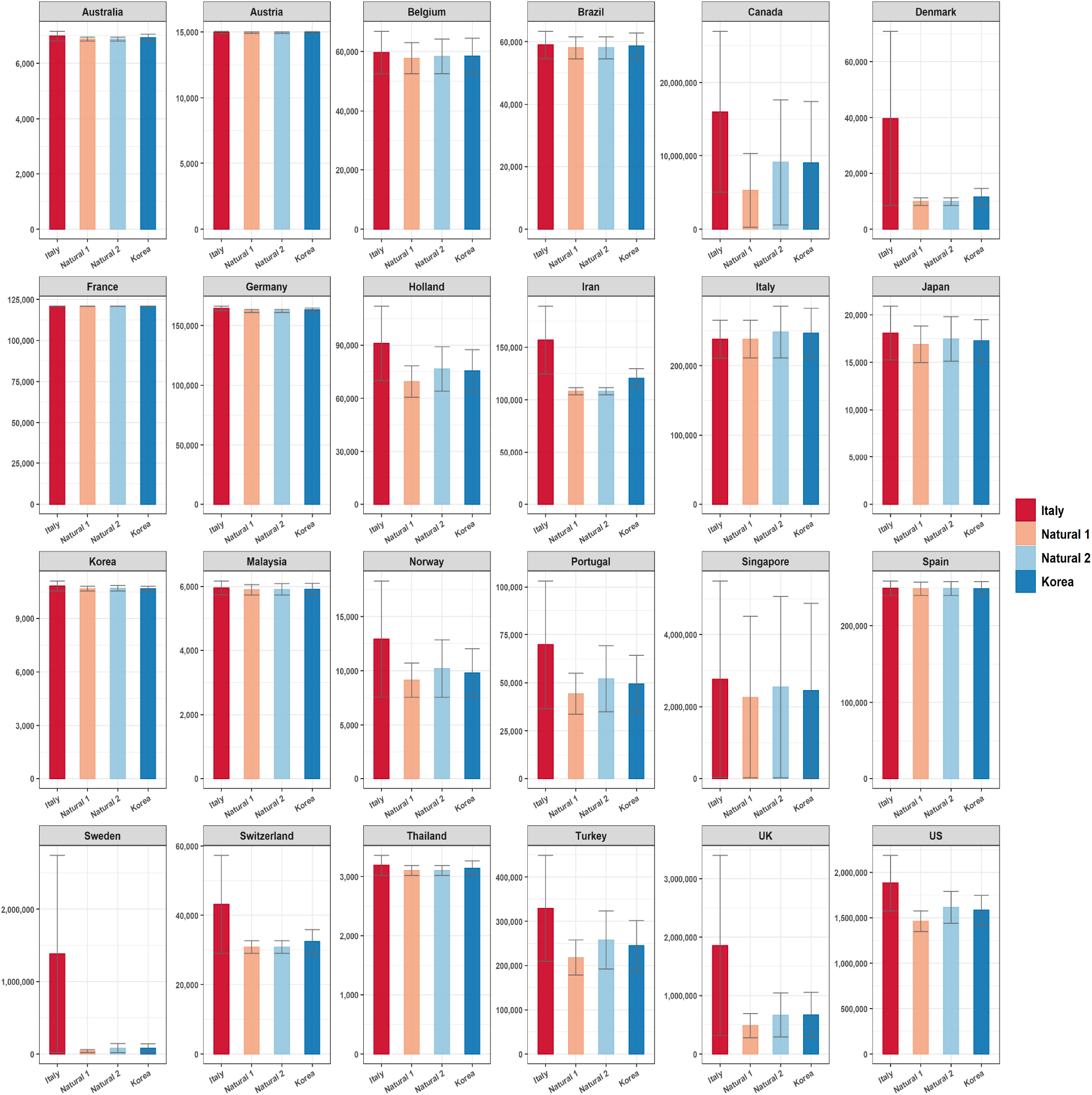
Projected total infected cases and the 95% prediction intervals under the two Natural Designs of respective countries, and Korea (blue) and Italy (red)’s recovery and death rates. Natural Designs 1 and 2 impose a minimum level of 1/17.5 (orange) and 1/28 (light blue) on the recovery rate of each country, respectively. The corresponding numerical results are provided in Table S9 of SM. The data used for the forecasting are up to April 20.

## Methods

### Data

We consider 25 countries in our study as listed in Table 1, which have experienced COVID-19 with at least four weeks of established community infections. The daily records of infected, dead and recovered patients are obtained from Johns Hopkins University Center for Systems Science and Engineering (*8*) and WHO (*9*) for the 25 countries, supplemented by these nations’ official health ministry’s statistics and Dingxiangyuan Pneumonia website (*10*). The population sizes are from the United Nation (*31*).

### Time-varying coefficient SEIdR model

Let *S*(*t*), *E*(*t*), *I*(*t*) and *R*(*t*) be the counts of the susceptible, exposed, infected and removed persons in a country at time *t*, respectively, where *R*(*t*) is the sum of the recovered *R_r_* (*t*) and the death *R_d_*(*t*). We propose an extended varying coefficient Susceptible-Exposed-Infected-Recovered (vSEIdR) model, which extends the SIR (*11*) and SEIR models (*14*) by allowing infections in the exposed *E* state. This would better capture the COVID-19 reality that most infections are made before clinical diagnosed.

The vSEIdR model has the following differential equations for the conditional Poisson means of the daily increments given the observation prior to date *t*:

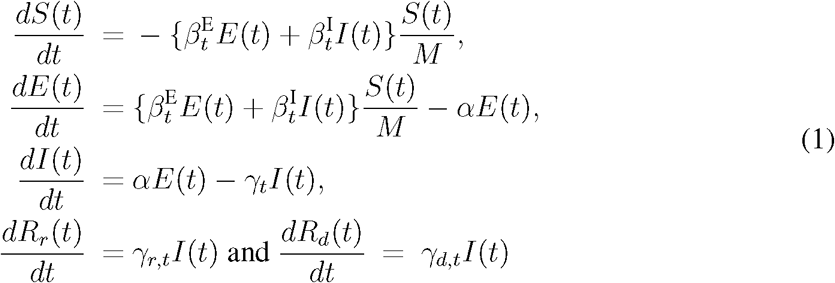

where 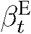, 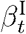, *γ_r,t_* and *γ_d,t_* are time-varying functions, representing the infection rates of the unconfirmed exposure group and the confirmed infected group, the recovery rate and the death rate, respectively, and *γ_t_* = *γ_d,t_* + *γ_r,t_* is the total removal rate. The diagnostic rate *α* is regarded as a constant, though extension to make it varying can be readily made.

The effective reproduction number under vSEIdR is

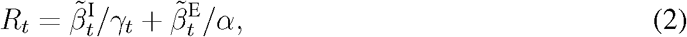

where 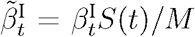 and 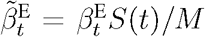, where *M* is the population size. As quite some countries have severe under-reporting in the recovered cases as reflected in the unseemly high proportion of death in the total removal rate in Table S6 in SM, we take *γ_t_* = 1/14 in calculating *R_t_* based on the clinical results for COVID-19 (*16*).

### Estimation

The reported numbers of confirmed infections are subject to measurement errors. To reduce the errors, we apply a weighted average with boundary kernel on the daily new infection number and new recovery number. Our estimation is based on the smoothed data. We assume the infectious rate 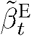 in the E-state is *r*-times of that in the I-state for a fixed constant *r* > 1 and all *t*. We first estimate 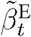, *γ_r_,_t_* and *γ_d_,_t_* under a fixed diagnosis rate *α*. The estimation of *α* is attained via the leave-one-out cross validation based on the fitting performance. See the SM for more details.

Let *N*(*t*) = *I*(*t*) + *R*(*t*) be the total number of accumulative infections at time *t*. Let Δ*S*(*t*), Δ*E*(*t*), Δ*N*(*t*), Δ*R_r_*(*t*) and Δ*R_d_*(*t*) be the daily changes in the compartments. From the progression of the epidemic in (1), we have the daily increase of new exposed and confirmed infections as

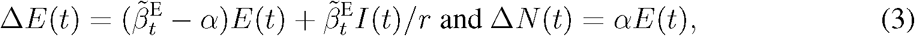

respectively. This implies we can estimate 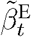 by a kernel weighted local regression of *E*(*t* + 1) + (*α* − 1)*E*(*t*) on *E*(*t*) + *I*(*t*)/*r* with imputed values for the unobserved numbers of the pre-diagnosed infections *E*(*t*). Similarly, *γ_r_,_t_* and *γ_d_,_t_* can be estimated by a local weighted regression of Δ*R_r_*(*t*) and Δ*R_d_*(*t*) on *I*(*t*), respectively. To conduct statistical inference for the estimates, we generate parametric bootstrap processes under the stochastic vSEIdR model with Poisson increment and the estimated parameters from the original data. The parameters 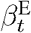, 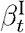, *γ_r_,_t_* and *γ_d_,_t_* are re-estimated using the bootstrap resampled data, and their confidence intervals are constructed based on the re-centered bootstrap estimates. See SM for more details.

### Start and Action Dates

The start date for established community transmission of a country is determined by considering the time when new confirmed cases of local infection emerges, the start date provided by the WHO for local infection (*9*) and the estimated infection rate under the vSEIdR model. The start date is determined by the first local maximum of the estimated infection rate after the WHO date.

The action dates for COVID-19 control measures are provided in Table S1 based on both governmental and credible media sources. When a series of measures are implemented over a time window, the middle date of the time window is used as the action time.

### Designs for scenario analyses

We project the numbers of confirmed infections and death under the Korea and China scenarios and earlier and delayed interventions for US and UK. The projections are made based on the hypothetical infectious rate under those scenarios and the proposed vSEIdR model, while keeping each country’s other epidemic parameters: the diagnose rate *α*, the recovery and death rates *γ_r_,_t_* and the *γ_d_,_t_* the same. The hypothetical infectious rates are constructed by applying the daily change of the estimated infectious rates of Korea and China from the 8th day since their starts of steady community transmission (Korea and China’s scenarios), or those of the UK and the US since their interventions to the target dates of scenario analysis. Figures S1, S2 in SM and Figure 4 display the projected infection rates and the effective reproduction numbers *R_t_* under those scenarios.

### Prediction

The prediction for the infection rates 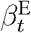 and 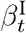 is attained by fitting the reciprocal model

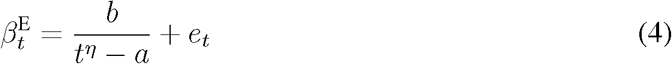

on their empirical estimates of the last 7 days before April 20, where *a, b* and *η* are unknown parameters and *e_t_* is an error term, and then projecting with the fitted parameters. The death and recovery rates *γ_d_,_t_* and *γ_r_,_t_* are set as the averages of the immediate past 7 days, and the diagnosis rate *α* is kept as the empirical estimates based on the whole data. See Table S6 for their estimated values.

For long term prediction, due to the under-reporting of recovery numbers in many countries as shown in Table S6, we set the long term removal rate for each country to be the maximum of their empirical estimates and *γ_r_*,_min_, where *γ_r_*,_min_ = 1/17.5 or 1/28 based on the clinical results (*29*,*30*) with a two week transitional period. We call the above settings as the Natural Designs 1 and 2 for each country, respectively. In all forecasting designs, each country’s projected infectious rate is used. The future projection of the epidemic is made based on the epidemic progression given in equations (1). Detailed procedures are provided in SM.

To assess the forecasting performance of the proposed model, we consider short term prediction with empirical removal rates for new infections in the next 7 days after April 13 and the next 14 days after April 6. Table S7 and Figure S4 in SM report the 7-day and 14-day prediction errors for the 24 countries. We see that the error percentages for the number of new infections of the 7-day and 14-day predictions were generally small, around 11.8% and 17.1% respectively, which justify the vSEIdR model and indicates its utility in epidemic prediction.

## Discussion

Our study shows that both the sizes of infection and the deaths of COVID-19 are particularly responsive to early containment measures as verified not only by the results from Korea and China, but also by the potential outcomes generated under the counter-factual experiments for the US and the UK. The roles of the removal rates are also important in particular to the final death tolls, and less so for the infection sizes for countries having had their *R_t_* fell below 1. There are several critical lessons one can deduce from the 25 countries’ COVID-19 experiences, especially for countries who are about to experience the first wave of the epidemic. The first one is to take action as early as possible to reduce the contact rates so as to reduce the infection rates and the *R_t_*. Acting early can hugely impact the infection size and thus lessen demands on medical resources down the track, and eventually improve the removal processes for those infected. The second lesson is to maintain a certain level of the diagnostic rate to speed up the epidemic progression as favorably shown in Korea, and the US and the UK diagnosis-rate-exchange experiment. Finally, at any stage of the epidemic, improving the recovery rates is always effective in reducing the deaths and the infection size as well.

## Data Availability

All data included in this study are available upon request by contact with the corresponding author.

## Supplementary materials

Materials and Methods

Supplementary Text

Tables S1 to S10

Figures S1 to S6

